# SCU-Net: A deep learning method for segmentation and quantification of breast arterial calcifications on mammograms

**DOI:** 10.1101/2021.07.30.21261406

**Authors:** Xiaoyuan Guo, W Charles O’Neill, Brianna Vey, Tianen Christopher Yang, Thomas J Kim, Maryzeh Ghassemi, Ian Pan, Judy Wawira Gichoya, Hari Trivedi, Imon Banerjee

## Abstract

**Purpose:** Measurements of breast arterial calcifications (BAC) can offer a personalized, noninvasive approach to risk-stratify women for cardiovascular disease such as heart attack and stroke. We aim to detect and segment breast arterial calcifications in mammograms accurately and suggest novel measurements to quantify detected BAC for future clinical applications.

**Methods:** To separate BAC in mammograms, we propose a light-weight fine vessel segmentation method Simple Context U-Net (SCU-Net). Due to the large image size of mammograms, we adopt a patch-based way to train SCU-Net and obtain the final whole-image-size results by stitching patch-wise results together. To further quantify calcifications, we test five quantitative metrics to inspect the progression of BAC for subjects: Sum of Mask Probability Metric (𝒫ℳ), Sum of Mask Area Metric (𝒜ℳ), Sum of Mask Intensity Metric (𝒮ℐℳ), Sum of Mask Area with Threshold Intensity Metric (𝒯𝒜ℳ_*X*_) and Sum of Mask Intensity with Threshold **X** Metric (𝒯 𝒮ℐℳ_*X*_). Finally, we demonstrate the ability of the metrics to longitudinally measure calcifications in a group of 26 subjects and evaluate our quantification metrics compared to calcified voxels and calcium mass on breast CT for 10 subjects.

**Results:** Our segmentation results are compared with state-of-the-art network architectures based on recall, precision, accuracy, F1-score/Dice Score and Jaccard Index evaluation metrics and achieve corresponding values of 0.789, 0.708, 0.997, 0.729, and 0.581 for whole-image-size results. The quantification results all show *>*95% correlation between quantification measures on predicted masks of SCU-Net as compared to the groundtruth and measurement of calcification on breast CT. For the calcifications quantification measurement, our calcification volume (voxels) results yield *R*^2^-correlation values of 0.834, 0.843, 0.832, 0.798, and 0.800 for the 𝒫ℳ, 𝒜ℳ, 𝒮ℐℳ, 𝒯𝒜ℳ_100_, 𝒯 𝒮ℐℳ_100_ metrics, respectively; our calcium mass results yield comparable *R*^2^-correlation values of 0.866, 0.873, 0.840, 0.774, and 0.798 for the same metrics.

**Conclusions:** SCU-Net is a simple method to accurately segment arterial calcification retrospectively on routine mammograms. Quantification of the calcifications based on this segmentation in the retrospective cohort study has sufficient sensitivity to detect the normal progression over time and should be useful for future research and clinical applications.

## I. Introduction

Cardiovascular disease is a source of high morbidity and mortality in women^1^. One of the barriers to improving diagnosis outcomes is the lack of a simple, inexpensive, and reliable method for screening and for assessing efficacy of therapies. Vascular disease commonly manifests as arterial calcifications, which are typically assessed by computed tomography (CT) or CT angiography of the coronary arteries and aorta^2^. However, these tests are expensive, usually performed only in symptomatic patients, and associated with additional radiation exposure. Calcification also occurs in breast arteries and can be readily observed on screening mammograms. The prevalence of breast arterial calcifications (BAC) correlates with calcifications in other arteries and is associated with an increased risk of cardiovascular disease events^3,4,5,6^. We recently showed that quantification of BAC through manual measurements can more accurately stratify risk factors and provide a means to follow progression^7,8,9^.

Each year, more than 40M women over age 40 undergo screening mammography for breast cancer screening^6^. Automatic detection and quantification of BAC in these women may be helpful in identifying patients at high-risk for cardiovascular events and following progression of vascular calcifications without additional cost or radiation exposure ^10^. Stored digital mammograms over the past decade would also provide a vast dataset for robust retrospective research. Currently, there is no standardized method for accurate detection, segmentation and quantification of BAC on mammography, which limits the utility of this potential biomarker. There are many challenges in automated detection of BAC. First, BAC appear as slender, elongated regions of fragmented high pixel intensity on mammograms and typically represent fewer than 1% of a 4*K* × 3*K* image. Moreover, the narrow appearance and potential variable lengths make precise segmentation of BAC much more challenging compared to general segmentation tasks. Second, there is no standard strategy for acquiring groundtruth BAC segmentations due to the variations in vessel width, severity of calcifications along the vessel, and tortuous vessel paths. Third, the large image size (over 12MP) adds significant difficulty in image processing.

Although there have been a number of existing works relevant to breast arterial calcifications, few have focused on accurate segmentation. Sulam et al. ^11^ examined only prevalence and Abriele et al.^12^, Juan et al.^13^ and Hossain et al.^14^ all detected BAC with a patch-based method, but did not report detailed segmentation performance or quantification metrics. Since BAC segmentation can be considered as a type of semantic segmentation in the realm of general computer vision, current semantic segmentation models can be attempted for BAC segmentation. Generally, semantic segmentation models can be classified into two main categories: non-real-time and real-time segmentation models. Non real-time models such as U-Net^15^, SegNet^16^, DeepLabV3^17^ and LinkNet^18^ usually have complex architectures and a high number of trainable parameters. Thus, they may achieve high accuracy but are slow to train and deploy. By contrast, real-time semantic segmentation models including ERFNet^19^, ESNet^20^, FastSCNN^21^, ContextNet^22^, DABNet^23^, EDANet^24^, FPENet^25^, CGNet^26^ have fewer trainable parameters but can still attain comparable performance with the non-real-time models. At our institution, up to 250 screening mammograms are performed daily constituting approximately 1,000 images. In live clinical deployment, it would be advantageous that BAC detection and quantification occur in near real-time so that the results are available to the interpreting radiologist in case patient referral is needed. Therefore, segmentation models with a high number of trainable parameters (*e*.*g*., U-Net^15^ has 13,395,329 parameters) would be prohibitive in their inference times, and lightweight models would enable more clinically viable.

To address the challenges and fulfill the requirement of clinical application, we propose Simple Context U-Net (SCU-Net), an automated lightweight segmentation model, to segment BAC in mammograms in a patch-based way. SCU-Net offers comparable performance of the most popular current segmentation architectures with an order of magnitude fewer training parameters. It achieves this by taking advantage of both dilated convolution operations and skip connections to learn and fuse global features with low-level information efficiently while maintaining far fewer trainable parameters. We demonstrate the efficacy of SCU-Net by visually and quantitatively presenting our BAC segmentation results as compared to a series of popular semantic segmentation models. Furthermore, we present five novel metrics to quantify the severity of BAC within the segmentation mask, compare our quantification metrics to breast CT, and demonstrate the ability to track a longitudinal increase in BAC in a cohort of patients with 10 years of retrospective mammograms. Thus, SCU-Net model may serve as a potential research and clinical tool for early detection and risk stratification of cardiovascular disease for women.

## II. METHODS

### II.A. Preprocessing

Mammograms contain a wide variety of pixel intensities with varying breast shapes and proportions of breast tissue versus null background. Therefore, image pre-processing is critical to identify breast tissue and normalize the image to maximize the model performance. To this end, we first smooth the image using median filtering^27^ with a disk kernel of size 5 for cleaning the noise but also avoiding causing serious blurring. This was chosen empirically among the evaluated range of [5-20] based on visual evaluation during preliminary experiments. To extract breast tissue only, we erode and then dilate the breast images with a disk kernel (size is 10 in our experiment) to erase the scanner labels of mammograms such as view type (*i*.*e*., “RMLO” – right mediolateral oblique, “LMLO” – left mediolateral oblique, “RCC” – right craniocaudal, “LCC” – left craniocaudal). With the same setting, we dilate and then erode the binary mask to link together and smooth any nearby annotation segments, producing a continuous vessel mask. Finally, we enhance image contrast to maximize the difference between calcified vessel and background tissue. During training, we normalize input image patches with zero-means method to minimize the impact of variation contrast between vessels and background.

### II.B. Network architecture

To overcome the issue of large image sizes and the inability to downsample images without data loss, we propose Simple Context U-Net (SCU-Net), whose inputs are patches cropped at the highest resolution of mammography images. The architecture of SCU-Net is shown in Figure 1. All the feature sizes in the figure are presented same as our experimental settings. SCU-Net is a symmetric, U-shaped model, similar to U-Net ^15^. The model has input image patches with size of 3 × 512 × 512. ^1^ The original input is fed into three 3 × 3 convolutional layers. To preserve the original image information, the input patch is downsampled with scale factor of 1 and 2. The obtained two downsampled input features are in size of 3 × 256 × 256 and 3 × 128 × 128 corresponding to the second and third green additional inputs of Figure 1. These two downsampled inputs will be concatenated with later high-level features. Each concatenation is followed by BatchNormalization and Parametric ReLU operations, enabling smooth fusion of high-level information with low-level features. Feature fusing is important, but the surrounding context is also very helpful for semantic segmentation^26^. Inspired by CGNet^26^ and DilatedNet^28^, SCU-Net adopts two different dilated convolutional layers (Dconv1 and Dconv2 in Figure 1) to aggregate multi-scale contextual information. In the decoder arm of the network, the learned image features are upsampled with bilinear interpolation and then concatenated with the corresponding encoder features of the same size. “Up” in Figure 1 means upsampling layer. Two 3 × 3 convoultional layers follow each concatenation. In total, there are three upsampling layers to get the network back to the original size. Finally, two 3 × 3 convolutional layers helps reduce the channel numbers to the class number, 1 in our case, and a Sigmoid layer is used to get the final mask prediction. All the convolutional layers including conv, Dconv1, Dconv2 and Up layers in Figure 1 are followed with BatchNormalization and Parametric ReLU operations. To avoid overfitting, we use online data augmentation techniques during training, including randomly vertical or horizontal flipping, randomly rotation by 90 or 270 degrees, and randomly changing the brightness, contrast and saturation of image.

**Figure 1:**
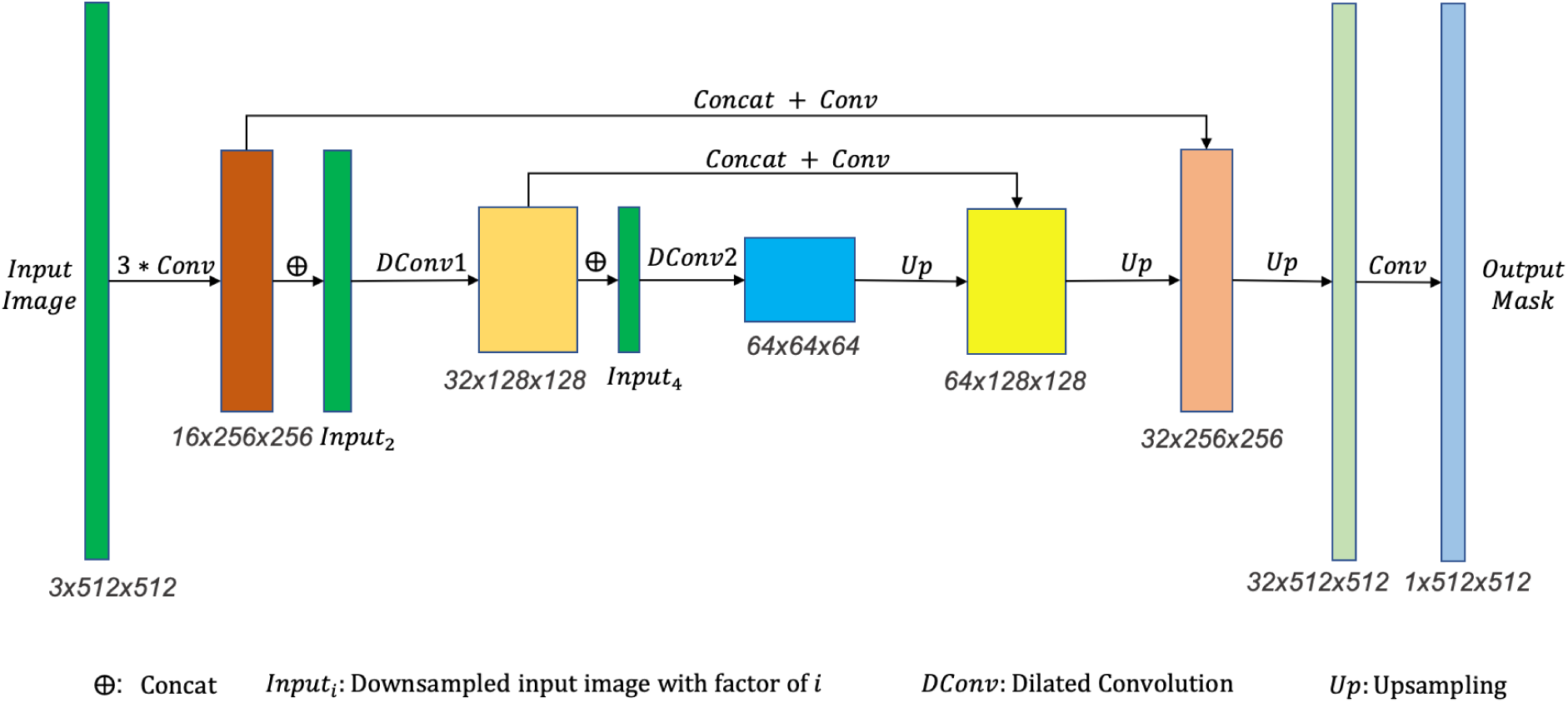
Network architecture of SCU-Net.

#### Implementation details

In our experiments, binary cross entropy loss converges much more slowly than dice loss, therefore we adopt dice loss to optimize all the segmentation networks. For optimization, we use Adamw optimizer with a learning rate of 0.001 for model training. Each network is trained with 50 epochs. The pipelines are developed using Pytorch 1.5.0, Python 3.0. and Cuda compilation tools V10.0.130 on a machine with 4 NVIDIA Quadro RTX 6000 with 24GB memory.

### II.C. Experimental setup

With the approval of Emory Institutional Review Board (IRB), three cohorts of subjects were identified from previous studies ^7,8,9^. All mammograms extracted were 2D full-field digital mammograms (FFDM) obtained during routine screening exams on Hologic (Marlborough, PA) mammography scanners in accordance with Mammography Quality Standards Act (MQSA) requirements. Screening exams consisted of four standard views - LCC, LMLO, RCC, RMLO.

- Cohort A – 661 FFDM from 216 subjects were annotated and used for deep learning model training and validation. The mean age was 70 ± 11 and 37% were African-American. Because the previous studies focused on kidney disease, 35% had chronic kidney disease, end-stage renal disease (ESRD), or renal transplantation. Mean breast density was 2.23 ± 0.77 as reported according to Breast Imaging Reporting and Data System (BI-RADS) guidelines (A=1, B=2, C=3, D=4). The majority of patients were density B (scattered fibroglandular tissue - 43.6%) and C (heterogeneously dense - 41.7%) with a minority of density A (mostly fat - 7.2%) and D (extremely dense - 7.5%).
- Cohort B for comparison to breast CT calcification - A previously reported cohort of 10 subjects with contemporaneous measurement of BAC by breast CT. Mean age was 69 ± 11 and all but one were Caucasian. Mean breast density was slightly lower at 2.08 ± 0.76.
- Cohort C for longitudinal analysis - 26 additional subjects with BAC and at least 5 yearly mammograms were studied in order to assess the ability to detect progression of BAC. The mean age was 65 ± 12 and 54% were African-American. Of these, 9 had ESRD or had undergone kidney transplantation. Mean breast density was similar at 2.19 ± 0.70.

#### Groundtruth acquisition

Mammograms from Cohort A were annotated by four annotators - one physician (CO) with 15 years experience and three other annotators trained and monitored by CO. Groundtruth segmentations are performed manually on whole images using the online platform Md.ai^2^ and standardized by annotating a multi-segmented line down the center of any calcified vessel continuously until there is at least a 1cm length of non-calcified vessel, at which point a new segmentation is started where the calcification resumes. These annotations serve as groundtruth training and validation data.

#### Data preparation

To prepare high-quality datasets for training deep learning models, the whole mammogram dataset is randomly divided into training and validation parts with 527 mammography images for training and 134 for validation. The mammography images are either sized 4096 × 3328 pixels or 3328 × 2560 pixels, which require a large amount of memory to load and analyze. Therefore, we crop images into fixed-size patches of 512 × 512 with 64 pixels of overlap between adjacent patches. The overlapping ensures the ability to connect BAC segmentations from adjacent patches and improves the overall segmentation accuracy. We exclude black background image patches to eliminate unnecessary calculations. Moreover, only patches that contain calcifications are left for segmentation training given the fact that the calcification mask prediction is pixelwise classification. Ultimately, this yields 3,455 effective patches for training and 901 patches for validation.

#### Model comparison

Experiments are performed with SCU-Net and state-of-the-art deep learning models including SegNet^16^, DeepLabV3^17^, U-Net^15^, LinkNet^18^, ERFNet^19^, ESNet^20^, FastSCNN^21^, ContextNet^22^, DABNet^23^, EDANet^24^, FPENet^25^ and CGNet^26^. Their number of trainable parameters, including SCU-Net, are compared in Figure 2. The larger the circle area for a model is, the more parameters the model contains. As can be seen, SegNet^16^ has the most parameters while FPENet^25^ contains the least. Our model, SCU-Net, has the second fewest parameters (marked in blue). Models with fewer parameters have lower complexity, consume less memory, and achieve faster training. Since mammograms (along with most radiology images) are very large in size, the number of model parameters is an important factor for real-world implementation as it is directly related to speed.

**Figure 2:**
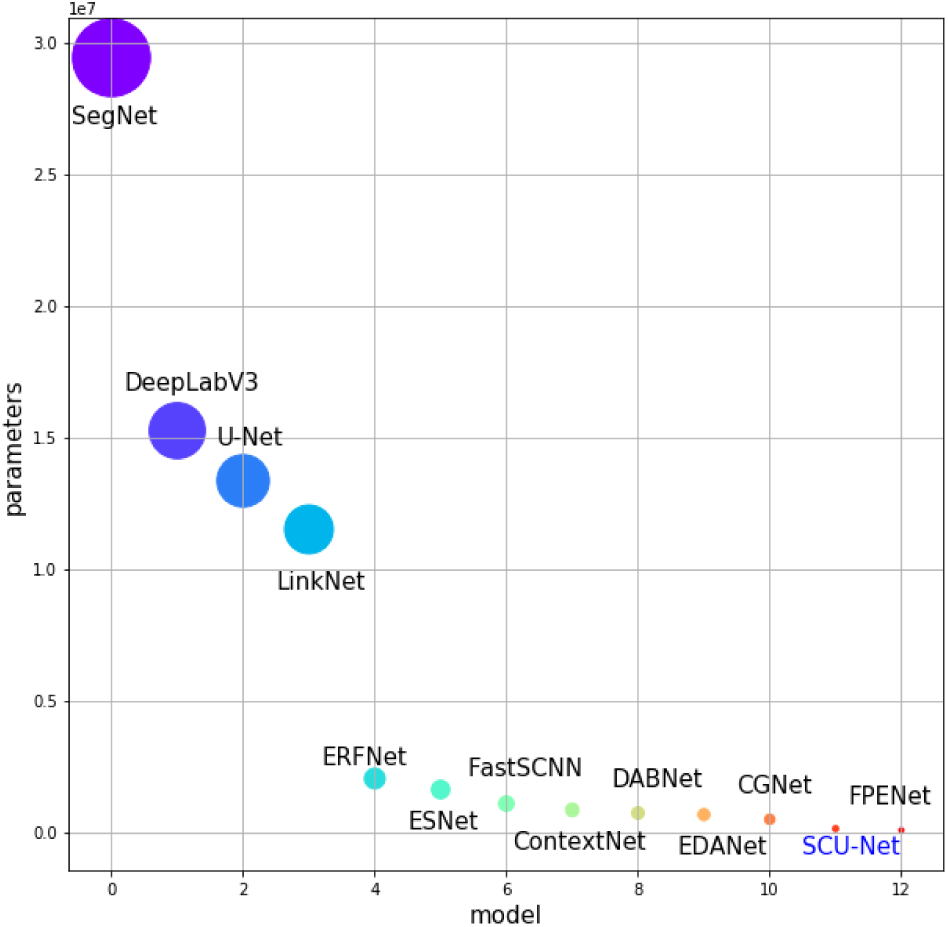
Trainable parameters comparison of segmentation models. The circle area is proportional to the total parameters of the model. Comparatively, SCU-Net is roughly two orders of magnitude smaller than other models.

#### Evaluation metrics for BAC segmentation

We evaluate both patch-wise segmentation results and final whole image segmentation results of all the models with five metrics: *Recall, Precision, Accuracy, F1-score/Dice score, Jaccard Index* value. The definitions are shown in Equations 1 and 2. In the equations, *TP, FN, TN* and *FP* calculations refer to pixelwise results.

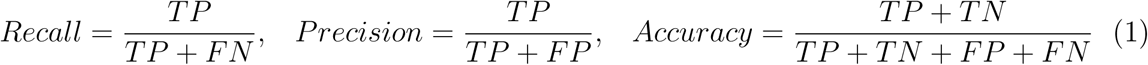

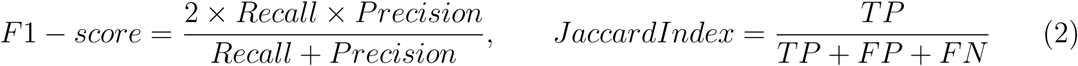

To further demonstrate the differences across all the models, we also perform pairwise t-test to compute the statistical significance of state-of-the-art models compared with SCU-Net. The p-value table is present in the supplementary material.

#### Evaluation metrics for BAC quantification

Beyond typical semantic segmentation evaluation metrics (*Recall, Precision, Accuracy, F1-Score/Dice Score and Jaccard Index*), we propose five BAC quantification metrics in Equations 3 and 4 to further measure the effectiveness of BAC detection in the predicted segmentation masks. Because of the segmentation challenges with BAC, we anticipated acceptable but imperfect segmentation results. However, unlike cancer detection where localization is extremely important, vessel segmentation can be considered an intermediate task to achieve BAC quantification. Slight differences in vessel segmentation region or width may have strong negative effects on standard evaluation metrics like Dice score and Jaccard index, but may still provide excellent results in terms of capturing clinically relevant calcifications. Therefore, we developed the following five metrics to capture the total segmented area, intensities of pixels within the segmented area, and thresholded pixel intensities and counts within the segmented area. Equations 3 and 4 show the definitions for Sum of Mask Probability Metric (𝒫ℳ), Sum of Mask Area Metric (𝒜ℳ), Sum of Mask Intensity Metric (𝒮ℐℳ), Sum of Mask Area with Threshold Intensity **X** Metric (𝒯𝒜ℳ_*X*_) and Sum of Mask with Intensity Threshold **X** Metric (𝒯 𝒮ℐℳ_*X*_). In the equations, *m* and *n* refer to the width and height of the mammogram, *p*_*i,j*_ is the probability value at *< i, j >* returned by the trained model, ℐ_*i,j*_ means the intensity value of pixel at *< i, j >* and **X** is the intensity threshold.

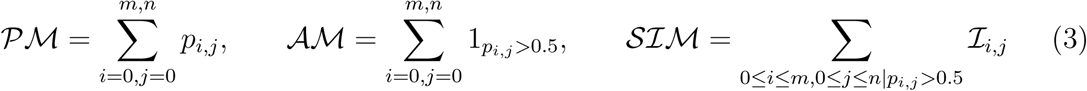

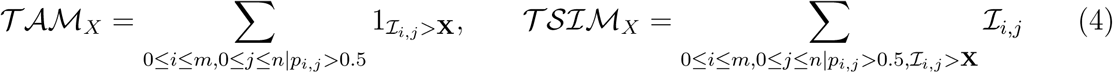

Specifically, 𝒫ℳ summates all predicted probabilities for an image to evaluate the confidence of the model’s prediction; 𝒜ℳ is the total number of pixels that are classified as BAC in a mammogram; 𝒮ℐℳ is the sum of the intensities of the pixels classified as BAC; 𝒯𝒜ℳ_*X*_ is the total number of BAC-classified pixels greater than intensity threshold **X**, as the BAC pixels usually have higher intensity values than background tissue area; 𝒯 𝒮ℐℳ_*X*_ is the sum of intensities for BAC-classified pixels with intensity value greater than the threshold **X**. In our experiment, we set **X** to be 100 as the best threshold for 𝒯𝒜ℳ_*X*_ and 𝒯 𝒮ℐℳ_*X*_ metrics based on visual observations of threshold values of 50, 75, 100, 150, 200. Metrics 𝒜ℳ, 𝒮ℐℳ, 𝒯𝒜ℳ_*X*_, and 𝒯 𝒮ℐℳ_*X*_ are all calculated with a model prediction cutoff of *p >* 0.5.

#### Comparison of BAC quantification metrics against breast CT measurements

To compare our quantification with a previously clinically validated measurement system^9^, we evaluated our quantification metrics on mammograms of 10 patients in Cohort B who had contemporaneous breast CT exams. All BAC quantification metrics on mammograms were compared to calcified voxels and calcium mass as measured on breast CT.

#### Evaluation of BAC quantification metrics longitudinally

To evaluate the utility of BAC quantification metrics to track calcification longitudinally, we examined 26 new subjects (Cohort C) not included in the original dataset with serial mammograms. Each patient had 5∼12 years imaging history with all four standard screening mammography views per exams, totalling 961 images across all subjects. SCU-Net was applied to each image to obtain the segmentation masks and 𝒯𝒜ℳ_100_ was calculated (based on top-performing correlation as shown in Figure 5). Plotting 𝒯𝒜ℳ_100_ over time *per view* initially yielded very noisy results in which calcification quantity appeared to oscillate over time, which typically would physiologically not occur. We then took the sum of the 𝒯𝒜ℳ_100_ for *all views* plotted against time, which somewhat decreased the fluctuation but did not eliminate it. Finally, we realized that each year the patient’s breast position and magnification of the mammogram could vary, meaning that the raw number of pixels as counted in the 𝒯𝒜ℳ_100_ metric would be dependent on breast magnification. To normalize for this effect, we took 𝒯𝒜ℳ_100_ metric divided by the breast area for each image and then sum this result across all four views. This was the final method used for longitudinal analysis.

## III. Results

### Evaluation of BAC detection based on standard metrics

Figure 3 shows the patchwise segmentation results of SCU-Net as compared to several semantic segmentation models including SegNet^16^, ContextNet^22^,U-Net^15^, CGNet^26^ and SCU-Net. The first row is of particular interest as it demonstrates ductal calcifications which are benign and unrelated to BAC, but can appear similar. SegNet^16^, ContextNet^22^, and U-Net^15^ each erroneously detect these ductal calcifications to varying degrees, however SCU-Net correctly ignores these. Interestingly, SCU-Net demonstrates similar performance to CGNet^26^ as they both utilize dilated convolution operations to learn context features. The second row of Figure 3 demonstrates a patch with overall lower image contrast and overlapping breast tissue which mimics linear calcifications. In this case, ContextNet^22^ detects the most false positive pixels. The third and fourth cases contain less noise and a clear difference from the background tissue, in which case all the models perform well at detecting BAC. In brief, image noise, low image contrast, and overlapping background tissue can all affect the quantitative accuracy of segmentation. The same types of errors are noticed on whole-image-size mask prediction as shown in Figure 4. For better visualization, only the breast region are kept by truncating the unnecessary background from the original mammograms. In this figure, the dice scores for the predicted masks of each case are labelled in the top right corner. As can be seen, overall performance for BAC segmentation is quite good although each model suffers from varying degrees of false positives due to issues with image noise, tissue contrast, and lookalike findings. We also see that some images are intrinsically more difficult with lower dice scores across the board for rows 1 and 2 in as compared to rows 3 and 4 in Figure 4. In general, the segmentation masks of ContextNet^22^ contain more false positive fragments than other results. Nevertheless, most of the BAC is captured by all the models. Notably, SCU-Net achieves comparable dice scores compared to SegNet^16^, U-Net^15^ and CGNet^26^ despite significantly fewer parameters.

**Figure 3:**
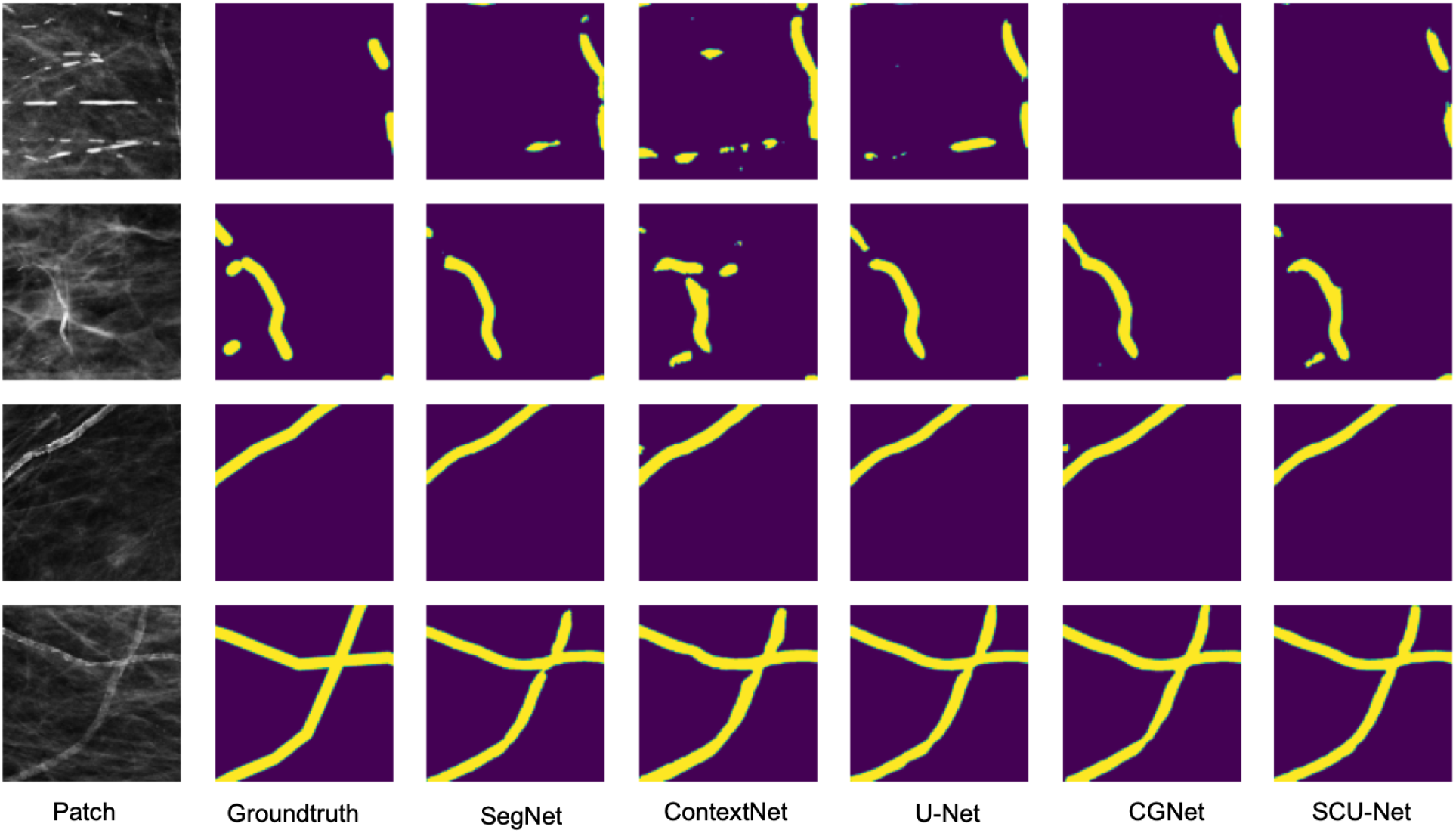
Examples of patch-wise segmentation results for BAC across multiple architectures as compared to the groundtruth. From left to right: original image patches, groundtruth mask, and prediction results of SegNet, ContextNet, U-Net, CGNet and SCU-Net.

**Figure 4:**
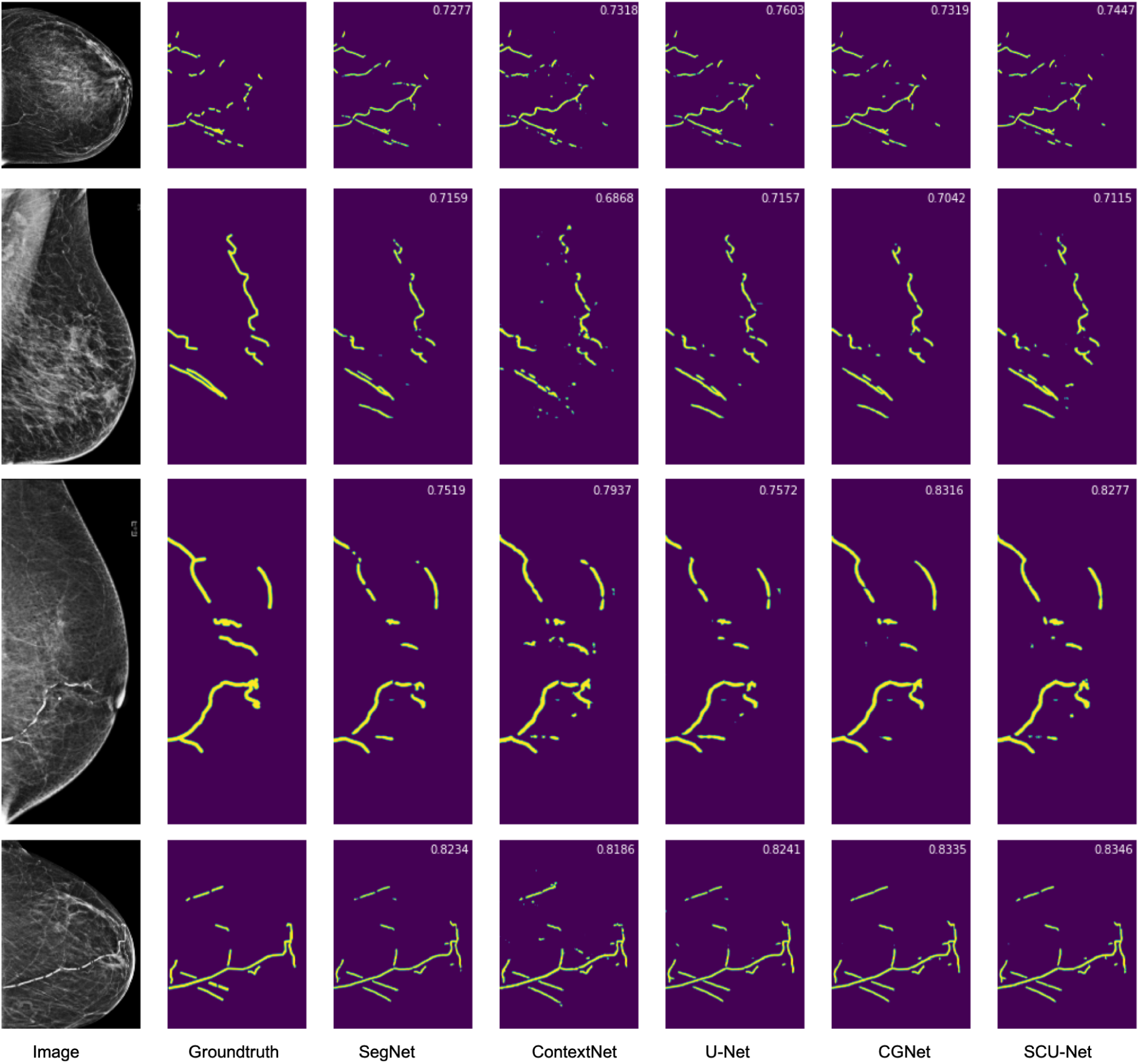
Examples of whole image segmentation results for BAC across multiple architectures as compared to groundtruth. From left to right: original mammography images (cropped to exclude background), groundtruth mask, prediction results of SegNet, ContextNet, U-Net, CGNet and SCU-Net. The F1-Score for each model is shown in the top right of the predicted mask. Higher F1-score means more overlap between groundtruth and the predicted mask.

**Figure 5:**
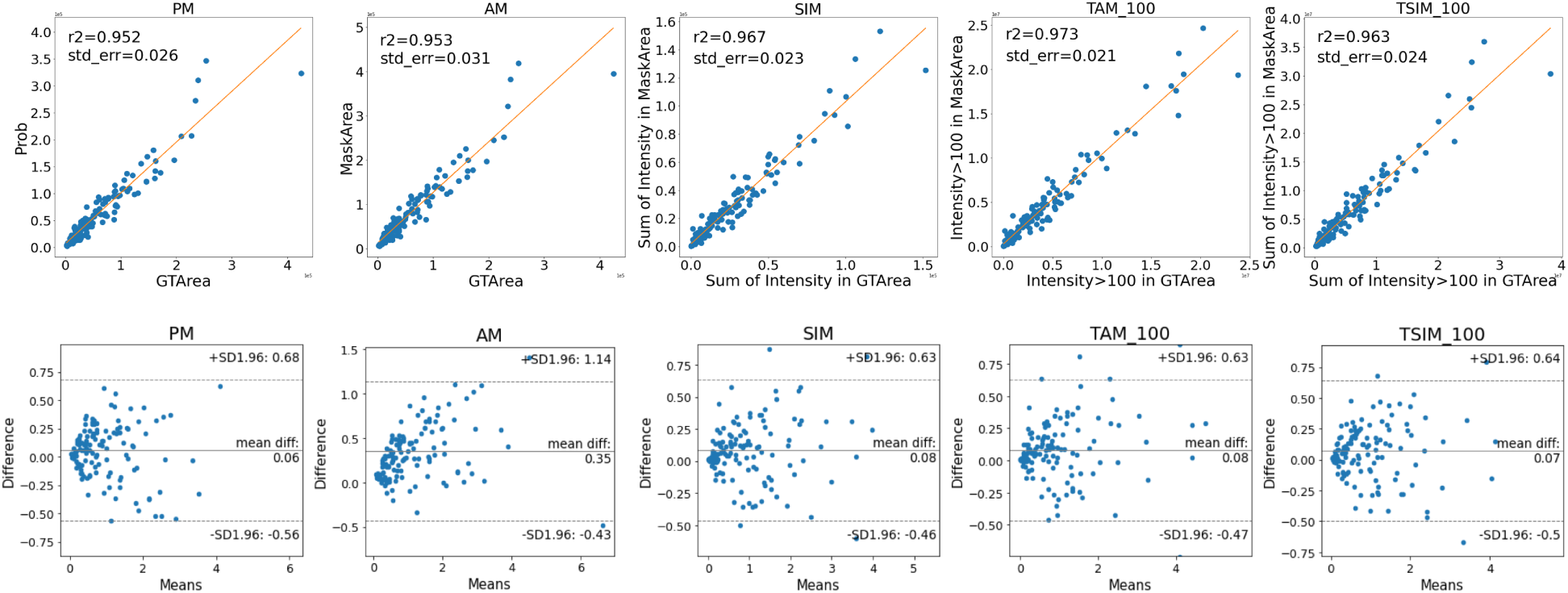
Statistical analysis on validation data for Cohort A. First row: *R*^2^-correlation of whole-image SCU-Net calcification quantification results for predicted masks (Y-axis) as compared to the groundtruth (X-axis). All X-axis and Y-axis values are in scientific format. *R*^2^-correlation values (r2) and standard errors (std err) are also reported for each metric in each subfigure. Second row: Bland Altman test to compare each metric computed from SCU-Net against the groundtruth. There are 134 data elements in total for each subfigure, with each point representing one image in the validation dataset.

Furthermore, we evaluate the segmentation results for both patches and whole images to demonstrate the fine vessel calcification segmentation accuracy. Table 1 presents the quantitative performance metrics for all tested models including SCU-Net, for both invidual patches (columns with clear background) and whole mammography images (columns with gray background). For patch-wise quantitative results in Table 1, ERFNet^19^ has the highest recall value, FPENet^25^ achieves the best precision value, and SCU-Net has the best F1-score and ties with CGNet^26^ for top Jaccard Index value. Accuracy values of all the models are relatively similar due to the high number of negative pixels in the image. Whole-image-size results are generated by concatenating the corresponding patches for each whole mammogram. Compared with patch-wise results, nearly all the evaluation metrics for the whole image are higher and are tightly grouped across all models. The reason lies in the overlapping 64 pixels with neighboring patches which helps enhance the segmentation accuracy by avoiding boundary effects^3^. On whole images, ERFNet^19^, FPENet^25^, DeepLabV3^17^ still maintain their advantages in recall, precision and accuracy respectively. U-Net^15^ and DeepLabV3^17^ in Table 1 have the best F1-score/Dice-score (0.735) and Jaccard Index value (0.59) for full image segmentation. With many fewer parameters (79x less), SCU-Net also performs very well with 0.729 of F1-score and 0.581 of Jaccard Index value compared with SegNet^16^ and FPENet^25^.

**Table 1:** Quantitative evaluation results for **image patches** (columns without background) and **whole images** (columns with gray background) in the validation dataset, subscripts denote standard deviation.

| Method | *Recall* |  | *Precision* |  | *Accuracy* |  | *F1-score* |  | *Jaccard* |  |
| --- | --- | --- | --- | --- | --- | --- | --- | --- | --- | --- |
| SegNet<sup>16</sup> | 0.707±0.100 | 0.764±0.159 | 0.704±0.095 | 0.743±0.128 | **0.981±0.005** | **0.998±0.002** | 0.676±0.084 | 0.734±0.098 | 0.554±0.079 | 0.589±0.113 |
| DeepLabV3<sup>17</sup> | 0.742±0.099 | 0.781±0.154 | 0.709±0.088 | 0.726±0.134 | **0.981±0.005** | **0.998±0.002** | 0.692±0.084 | **0.735±0.100** | 0.568±0.081 | **0.590±0.118** |
| U-Net<sup>15</sup> | 0.738±0.092 | 0.789±0.144 | 0.704±0.088 | 0.723±0.141 | **0.981±0.005** | **0.998±0.002** | 0.689±0.074 | **0.735±0.097** | 0.562±0.073 | **0.590±0.112** |
| LinkNet<sup>18</sup> | 0.748±0.095 | 0.801±0.151 | 0.675±0.096 | 0.690±0.137 | 0.979±0.006 | 0.997±0.002 | 0.676±0.082 | 0.720±0.101 | 0.550±0.080 | 0.572±0.114 |
| ERFNet<sup>19</sup> | **0.788±0.088** | **0.826±0.133** | 0.669±0.086 | 0.673±0.151 | 0.979±0.006 | 0.997±0.002 | 0.694±0.075 | 0.724±0.106 | 0.568±0.077 | 0.578±0.123 |
| ESNet<sup>20</sup> | 0.757±0.096 | 0.796±0.164 | 0.684±0.091 | 0.707±0.137 | 0.980±0.005 | 0.997±0.002 | 0.687±0.083 | 0.727±0.108 | 0.563±0.081 | 0.581±0.122 |
| FastSCNN<sup>21</sup> | 0.687±0.105 | 0.738±0.171 | 0.662±0.100 | 0.695±0.136 | 0.979±0.006 | 0.997±0.002 | 0.647±0.096 | 0.697±0.112 | 0.522±0.092 | 0.545±0.124 |
| ContextNet<sup>22</sup> | 0.723±0.093 | 0.765±0.165 | 0.631±0.090 | 0.628±0.150 | 0.977±0.006 | 0.997±0.003 | 0.643±0.083 | 0.671±0.123 | 0.509±0.081 | 0.517±0.130 |
| DABNet<sup>23</sup> | 0.750±0.096 | 0.804±0.143 | 0.692±0.095 | 0.706±0.142 | **0.981±0.005** | **0.998±0.002** | 0.686±0.082 | 0.734±0.102 | 0.564±0.079 | 0.589±0.118 |
| EDANet<sup>24</sup> | 0.771±0.094 | 0.810±0.150 | 0.666±0.096 | 0.682±0.137 | 0.980±0.005 | 0.997±0.002 | 0.685±0.085 | 0.723±0.102 | 0.559±0.083 | 0.575±0.117 |
| CGNet<sup>26</sup> | 0.766±0.090 | 0.798±0.149 | 0.689±0.087 | 0.703±0.138 | 0.980±0.005 | 0.997±0.002 | 0.696±0.074 | 0.730±0.102 | **0.569±0.075** | 0.584±0.118 |
| **SCU-Net** | 0.778±0.085 | 0.789±0.137 | 0.682±0.082 | 0.708±0.140 | 0.980±0.005 | 0.997±0.002 | **0.698±0.074** | 0.729±0.093 | **0.569±0.074** | 0.581±0.110 |
| FPENet<sup>25</sup> | 0.682±0.106 | 0.730±0.173 | **0.715±0.095** | **0.750±0.130** | **0.981±0.005** | **0.998±0.002** | 0.666±0.092 | 0.721±0.114 | 0.544±0.087 | 0.575±0.129 |

### Evaluation of BAC quantification based on defined metrics

Universal semantic segmentation evaluation metrics are helpful in evaluating segmentation results by performing pixel-to-pixel evaluation. However, the ultimate goal of this work is to quantify the amount of BAC within a mammogram for eventual correlation with cardiovascular outcomes. To evaluate the practical performance of SCU-Net’s segmentations in capturing BAC, we computed the correlation for all metrics computed using SCU-Net segmentations against the same metrics computed on the ground truth segmentation. The upper row of Figure 5 shows the *R*^2^-correlation of whole-image-size segmentation results of SCU-Net compared to the groundtruth based on the same metrics, demonstrating correlation *>*0.95 for all metrics. On the 134 validation scans, SCU-Net has the highest *R*^2^-correlation value of 0.973 between the predicted mask and groundtruth when using the 𝒯𝒜ℳ_100_ metric, which measures the total number of pixels with intensity *>*100 in the segmented mask. The second row of Figure 5 indicates the Bland Altman test results^29^ for the same validation data. The plots show the differences between quantitative metrics computed from the groundtruth and SCU-Net against the mean of the two measurements. Most metrics demonstrate very few outliers, and in particular 𝒫ℳ does not have a single outlier.

### Results of BAC quantification compared to breast CT

Evaluation of BAC quantification against breast CT in cohort B yielded good results. For calcification volume (voxels), *R*^2^-correlation values were 0.834, 0.843, 0.832, 0.798, and 0.800 for the 𝒫ℳ, 𝒜ℳ, 𝒮ℐℳ, 𝒯𝒜ℳ_100_, 𝒯 𝒮ℐℳ_100_ metrics, respectively. For calcium mass, *R*^2^-correlation values were comparable at 0.866, 0.873, 0.840, 0.774, and 0.798 for the same metrics. Although breast CT is not performed clinically, this demonstrates that BAC quantification on mammography is comparable to a previously validated calcification quantification metric.

### Results of BAC longitudinal analysis

Results of longitudinal analysis using the 𝒯𝒜ℳ_100_ metric showed the ability to automatically track BAC over time. Plots for five subjects shown in Figure 6 demonstrate a gradual increase in BAC over time. Figure 6 also shows five mammograms that demonstrate the progression of BAC in one subject over an 11 year period with predicted BAC masks highlighted in green.

**Figure 6:**
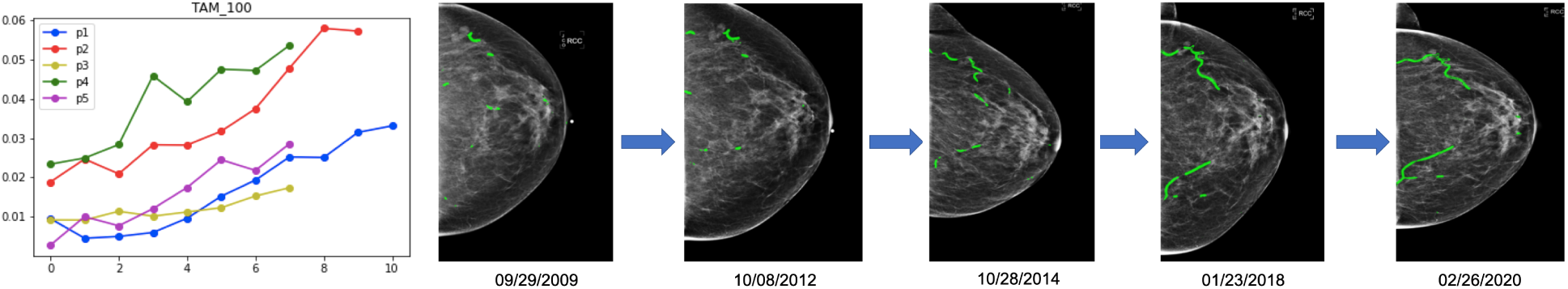
Longitudinal quantification of BAC in 5 patients. Left: The top-performing 𝒯𝒜ℳ_100_ metric applied to SCU-Net segmentations for five subjects plotted over time over time, wherein p1, p2, p3, p4, p5 represent different subjects. Right: Sampled mammograms from one subject over 11 years demonstrating an increase in detected BAC over time. BAC are highlighted in green. Each mammogram is cropped to exclude background with its exam date shown below.

## IV. DISCUSSION

We present a lightweight and accurate semantic segmentation model Simple Context U-Net (SCU-Net) designed for efficient vessel calcification segmentation on mammograms. It incorporates dilated convolution operations to learn context features and fuses multi-level features to enhance prediction accuracy. Due to the large size of mammograms, each image is processed in patches for both training and validation and the resultant masks are re-stitched to obtain whole-image predictions. Extensive experimental results for both patches and whole mammography images of 216 subjects showed comparable or better performance of SCU-Net as compared to current state-of-the-art models while maintaining far fewer training parameters. A further advantage of our model is that it does not require raw mammography data and can be applied retrospectively. This will enable analysis of the vast datasets of prior digital mammograms, allowing for large retrospective studies.

In addition to accurate segmentation of BAC, we applied quantification metrics to assess the extent of calcification and demonstrated excellent correlation between quantification values obtained on the predicted mask as compared to the groundtruth. Correlation was best using the 𝒯𝒜ℳ_100_ metric which counts all pixels above intensity 100 to differentiate between calcified and non-calcified portions of the vessel inside the mask. We also showed strong correlation of all metrics to calcium volume and mass obtained on breast CT for 10 subjects. Lastly, we were able to track and quantify the progression of BAC in 26 subjects longitudinally using this metric. Thus we believe this tool can accurately quantitatively measure and track BAC progression in patients and could be used to assess the efficacy of therapies and risk factors modification.

A limitation of this work is that the model is developed at a single institution using a single brand of scanners. It is possible that the model could underperform on external data, however we believe that the model can be successfully fine-tuned to re-optimized as needed, particularly due to its low number of parameters. The model is developed using only 661 images so fine-tuning can likely be achieved using an even smaller segmented dataset if needed. Another current limitation is that although our quantification metrics show strong correlation to breast CT data and track increases in BAC over time, they have not yet been validated against clinical outcomes in these patients. To address this in future work, we plan to evaluate our model and quantification metrics against outcomes data or existing validated risk assessment tools such as calcium scores on coronary CT.

In summary, a robust, minimally complex, deep learning method for segmenting and quantifying breast arterial calcifications has been developed that can be applied retrospectively to routine screening mammograms. This will allow for analysis of large populations without additional imaging costs or radiation exposure. Future studies will determine the performance of this tool for predicting clinical outcomes and determining the efficacy of prevention approaches.

## Data Availability

The datasets generated during and/or analyzed during the current study are not publicly available due to patient data privacy restrictions, but a de-identified subset of the data is available on reasonable request.

## VI. ACKNOWLEDGEMENTS

The work is supported by Winship Invest$ Pilot Grant Program. We thank Dr. Pradeeban Kathiravelu for helping with revision of manuscript. We also thank Dr. Anouk Stein and George Shih of MD.ai for providing their annotation platform.

## Footnotes

1 Although the mammogram image is grayscale and has only one image channel, three duplicates of the mammogram patch are stacked together to form a three-channel image same as RGB image format. This setting ensures the model to work for both natural and grayscale images, and can be comparable with existing segmentation models.

2 www.md.ai

3 Cropped patches may only contain a very small piece of calcification along the cropped boarder, which is hard to segment accurately. However, the larger calcification can be more easily detected in the adjacent patches. Thus, concatenating the predictions of adjacent patches can eliminate the boundary effects.

